# Establishing a physiological normative value of area-based hypoxic burden in obstructive sleep apnea

**DOI:** 10.64898/2026.07.31.26359352

**Authors:** Li Zhou, Siyun Yang, Sajila D. Wickramaratne, Pavel Boulgakov, Zachary Roberts, Sarah chu, Ankita kumar, Eriko Hamada, Thomas M. Tolbert, Korey Kam, Andrew W. Varga, Luciana B. M. de Godoy, Luciana O. Palombini, Monica L. Andersen, Sergio Tufik, Katie L. Stone, Indu Ayappa, David M. Rapoport, Ankit Parekh

## Abstract

**Background and Objectives:** Area-based hypoxic burden, termed as hypoxic dip area (HDA) has been studied as a novel obstructive sleep apnea (OSA) metric that may better characterize intermittent hypoxemia in OSA and thereby better predict outcomes. However, to be of use in routine clinical care, a physiological normative value is necessary. Here we attempt to use several large cohorts to establish a percentile-based threshold that can classify individuals with normal vs. abnormal physiological profiles.

**Methods:** Data from 10 cohorts (EPISONO, FINS, Dayfun, MrOS, MESA, SHHS, APPLES, WSC, CFS, and AIRS [Mount Sinai Clinical Cohort]; n=14,031 subjects) were included. HDA was defined as the area bounded by SpO_2_ nadirs (≥2% desaturation) flanking left/right peaks. The 97.5^th^ percentile of HDA among asymptomatic subjects (n=136) from EPISONO, FINS, and DAYFUN (aged 20-52 years; 46 [33.8%] male) was used to define normal and elevated HDA levels. Physiological features, clinical comorbidities, and incident cardiovascular disease (CVD) events and mortality were compared between normal/elevated HDA groups. Sensitivity analyses were conducted using age-, sex-, and BMI-adjusted threshold derived from multivariable regression model.

**Results:** The 97.5% percentile cutoff of HDA was 8.6%min/h, classifying 4,500 individuals as normal HDA and 9,531 as elevated HDA. Individuals with elevated HDA (≥8.6%min/h) demonstrated greater nocturnal hypoxemia (lower baseline SpO_2_, higher T90, T85, T80 and oxygen desaturation index), higher AHI (apnea-hypopnea index) and arousal index, and greater daytime sleepiness as well as a higher prevalence of lifetime CVD and hypertension across cohorts. Elevated HDA was related with a higher prevalence of incident CVD events and mortality in unadjusted analyses, although these associations were attenuated after adjustment for confounding factors. Results were similar using alternative thresholds derived from multivariable regression analyses.

**Conclusions:** A physiologically derived cutoff of 8.6% min/h for HDA effectively distinguishes physiological profiles across respiratory, arousal, and oxygenation domains and generalizes across multiple cohorts. Those with elevated HDA had a higher prevalence of incident CVD outcomes, although observed relationships were attenuated after further adjustment. Our findings provide a physiologically interpretable threshold for HDA that can be readily used in clinical use.

## Introduction

Obstructive sleep apnea (OSA) affects approximately one billion individuals worldwide and is associated with short- and long-term adverse outcomes, including excessive daytime sleepiness (EDS), cardio-cerebrovascular morbidity, and cognitive impairment.^1,2^ The pathophysiological changes in OSA are primarily driven by sleep fragmentation, intermittent hypoxemia, and recurrent arousals resulting from recurrent upper airway collapse during sleep.^3–5^ Currently, the diagnosis and severity classification of OSA rely primarily on the apnea–hypopnea index (AHI). However, AHI reflects only the frequency of apneic and hypopneic events per hour of sleep and has increasingly been recognized for its limitations in accurately representing disease severity, particularly with respect to the of hypoxemia.^6^

Characterizing intermittent hypoxemia in OSA, such as that done by hypoxic burden, has shown promise in terms of prognostic value and clinical utility.^7–10^ Broadly, hypoxic burden can be categorized into index-based measures (e.g., oxygen desaturation index [ODI3]), time-based measures (e.g., percentage time below SpO_2_ 90%, T90), and area-based measures, notably the hypoxic dip area (HDA), the latter of which have gained increasing attention in recent years.^7,10–15^ Index-based and time-based measures are limited to capturing either the depth or the duration of hypoxemia, whereas HDA incorporates both dimensions, indicating a potentially more comprehensive representation of hypoxemic severity.^15^ Prior evidence suggests that HDA metrics are more strongly correlated with short- and long-term outcomes compared with AHI, supporting their potential utility as surrogate markers for quantifying the severity of nocturnal hypoxemia.^8,10,12,16,17^

Several approaches have been proposed to calculate HDA, differing in their definitions of the reference baseline, including fixed 100% saturation, pre-specified thresholds, or event-specific levels (e.g., desaturation onset or local peaks).^8,10,16,18–20^ Many of these methods rely on manually annotated respiratory events, which are time-intensive and limit scalability.^10,16,18,19^ In contrast, our group has pioneered fully automated algorithms that eliminate the need for manual annotation.^8,13,20^ Studies alongside our prior work found that fully automated approach derived HDA are strongly connected with both short- and long-term outcomes, including EDS, incident cardiovascular disease (CVD), and long-term mortality, indicating their potential for clinical applicability and large-scale implementation.^8,13,21^

Despite these advances, there remains substantial heterogeneity in how HDA being used to stratify patients with different hypoxemia profiles. Existing studies commonly apply data-driven categorizations (e.g., median, tertiles and quintiles), which are highly dependent on the underlying sample distribution and may limit generalizability across populations and clinical settings.^8,10,16,22^ A recent study reported discordance between HDA and AHI-defined OSA severity, with higher HDA observed in mild-to-moderate OSA patients, but lower HDA in severe OSA patients.^23^ These findings suggest that AHI-defined OSA severity categories may not be appropriate for characterizing HDA, underscoring the need for HDA-specific cutoff values. Establishing normative, percentile-based thresholds for HDA using a standardized, fully automated approach may enhance its clinical utility for risk stratification in OSA.

The objectives of this study were to: (1) establish percentile-based cutoffs for HDA derived from a fully automated pipeline using a population of asymptomatic individuals; (2) evaluate the physiological relevance of these cutoffs by examining differences in respiratory and oxygenation characteristics across multiple community-based and clinical cohorts; and (3) compare the differences of clinical comorbidities and outcomes, particularly short-term EDS and long-term mortality, between normal and elevated HDA groups.

## Methods

### Study population

Our study used retrospective data from 10 cohorts. We used asymptomatic subjects’ data from 3 datasets: 1) Asymptomatic subjects from the epidemiological cohort (EPISONO [Sao Paulo Epidemiologic Study], which included 1,042 participants in total, with 106 classified as asymptomatic,^24,25^ 2) Asymptomatic subjects from an existing study (DAYFUN [Relating Sleep Disordered Breathing to Daytime Function], n=14),^26^ 3) Asymptomatic subjects from an existing Functional Imaging of Navigation across Sleep (FINS, n=16),^27^ to derive the HDA percentile-based cutoff. All asymptomatic subjects were free of known conditions that could affect respiratory function, gas exchange, or oxygenation (e.g., chronic pulmonary or cardiovascular diseases), as well as medications that may influence nocturnal oxygenation.

Additional 7 datasets were used, 6 from the National Sleep Research Resource (NSRR),^28,29^ Sleep Heart Health Study (SHHS, n=5,018),^30^ Osteoporotic Fractures in Men (MrOS, n=2,759),^31–33^ Multi-ethnic Study of Atherosclerosis (MESA, n=1,326),^34,35^ Cleveland Family Study (CFS, n=566),^36^ Wisconsin Sleep Cohort (WSC, n=1,037),^37,38^ and the Apnea Positive Pressure Long-term Efficacy Study (APPLES, n=1,062)^39,40^—and a real-world clinical dataset of patients with suspected OSA seen at the Mount Sinai Health System between 2016 and 2023 (Artificial Intelligence Research in Sleep Apnea [AIRS] Cohort, n=2,263), were used to evaluate the utility of the cutoff.

All subjects underwent overnight polysomnography (PSG) either at home or in the laboratory. Only subjects with valid data (defined as more than 3 hours of recording with valid oxygen saturation and respiratory effort signals) and good-quality SpO₂ signals were included in this study. Details for each dataset are provided in the supplementary materials.

### Derivation of Hypoxic Dip Area (HDA)

The SpO₂ signal was preprocessed to remove artifacts or any non-physiological segments of the signal. The details of automated HDA method were reported in our previous paper in supplementary methods.^8^ Desaturation events were automatically identified as decreases in SpO₂ of ≥2% lasting at least 3 seconds. A 2% threshold was selected based on prior evidence demonstrating that 2% desaturations are associated with EDS and long-term CVD mortality, whereas 3% and 4% thresholds were not.^13^ For each detected event, the preceding and subsequent local maxima were identified, and the area between these peaks and the nadir was calculated to quantify HDA, which was normalized by total recording time as described in prior study.^13^

### Outcomes

Respiratory events, arousal, and oxygenation profiles derived from PSG were used as physiological outcomes. EDS symptom outcome was defined as an Epworth Sleepiness Scale (ESS) score ≥10. For clinical outcomes, lifetime comorbidities, including CVD, hypertension, diabetes mellitus, etc, were assessed in the SHHS, WSC, CFS, and APPLES datasets. Incident CVD events (including angina, congestive heart failure, myocardial infarction, stroke, and coronary revascularization procedures) were evaluated in the SHHS, MrOS, and AIRS cohorts, with events in AIRS defined as the first incident CVD event using a combination of ICD and CPT. Mortality outcomes (all-cause and CVD) were assessed in the SHHS and MrOS datasets.

### Covariates

Age, sex, body mass index (BMI), race, smoking status, study cohort, total recording time, and ventilatory burden (VB) were as covariates. The details of VB derived method were shown in our previous paper.^8^

### Determination of the “normative” threshold for HDA

The 97.5^th^ percentile was used to define the upper reference limit of the distribution in asymptomatic individuals, a common approach for clinical reference interval estimation in sufficiently large data (n>120).^41,42^ Because a minimum value of 0 for HDA is considered normal, only the upper reference limit was defined, and a lower reference limit was not estimated. Individuals with HDA values below the 97.5^th^ percentile were characterized as those with normal HDA, whereas those with values above this threshold were identified as the elevated HDA group. To account for potential effects of demographic and anthropometric factors, sensitivity analyses were conducted using regression-based methods adjusted for age, sex, and BMI.

### Statistical analysis

Descriptive statistics are presented as median (interquartile range, IQR) for non-normally distributed continuous variables and as counts (percentages) for categorical variables. The standard Tukey IQR method was used to screen for potential outliers in HDA among asymptomatic individuals prior to deriving normative cutoff.^43^ Specifically, the first (Q1) and third (Q3) quartiles were calculated, and the IQR was defined as Q3– Q1. Observations exceeding Q3+1.5×IQR were considered potential outliers, given that a minimum value of 0 is considered normal. For these potential outliers, assessment of signal continuity, noise, and potential artifacts were checked to further determine whether they represented true outliers of physiologically impossible values.

To examine the relationships between elevated HDA (independent variable) and physiological as well as clinical outcomes (dependent variables), we performed regression analyses with and without covariates adjustment. Linear regression was used for continuous outcomes, logistic regression for binary outcomes (e.g., EDS and clinical comorbidities), and Cox proportional hazards regression for incident events. Regression models were fitted in combined cohort and separately within each study cohort. Sensitivity analyses applied alternative age-, sex-, and BMI-adjusted threshold derived from multivariable regression model. All analyses were conducted using SPSS (version 31.0.1.0) and R (version 4.5.1). A two-sided p value <0.05 was considered statistically significant.

## Results

### Cohort characteristics

Across the datasets with asymptomatic subjects, a total of 136 subjects were assessed, with ages ranging from 20 to 52 years (median 29.0 years), 33.8% were male, and the median BMI was 22.9 (Table 1). During an overnight PSG, subjects exhibited normal sleep patterns, with a median total sleep time of 361.0 (IQR: 90.3) minutes and sleep efficiency of 87.8% (IQR: 12.6). The median AHI3a was 0.5 (IQR: 1.0) events/h, and median HDA was 0.8%min/h (IQR: 1.9) and median T90: 0.0% (IQR: 0.0). The median ESS score was 6.0 (IQR: 7.0). An additional 14,031 subjects from 7 independent cohorts (SHHS, MrOS, MESA, WSC, CFS, APPLES, and AIRS) were analyzed for validation of the HDA cutoff (Table 1).

**Table 1.**
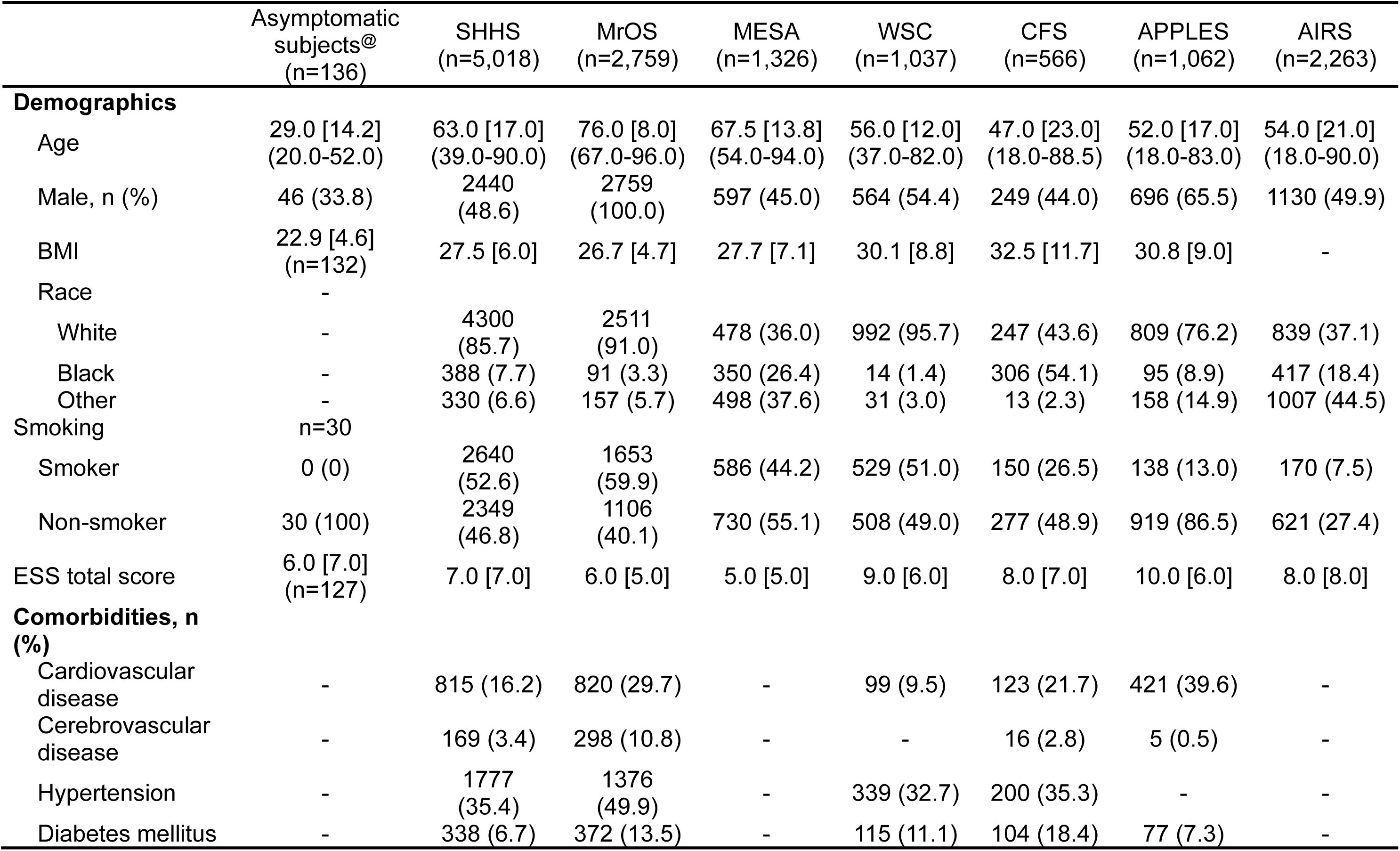
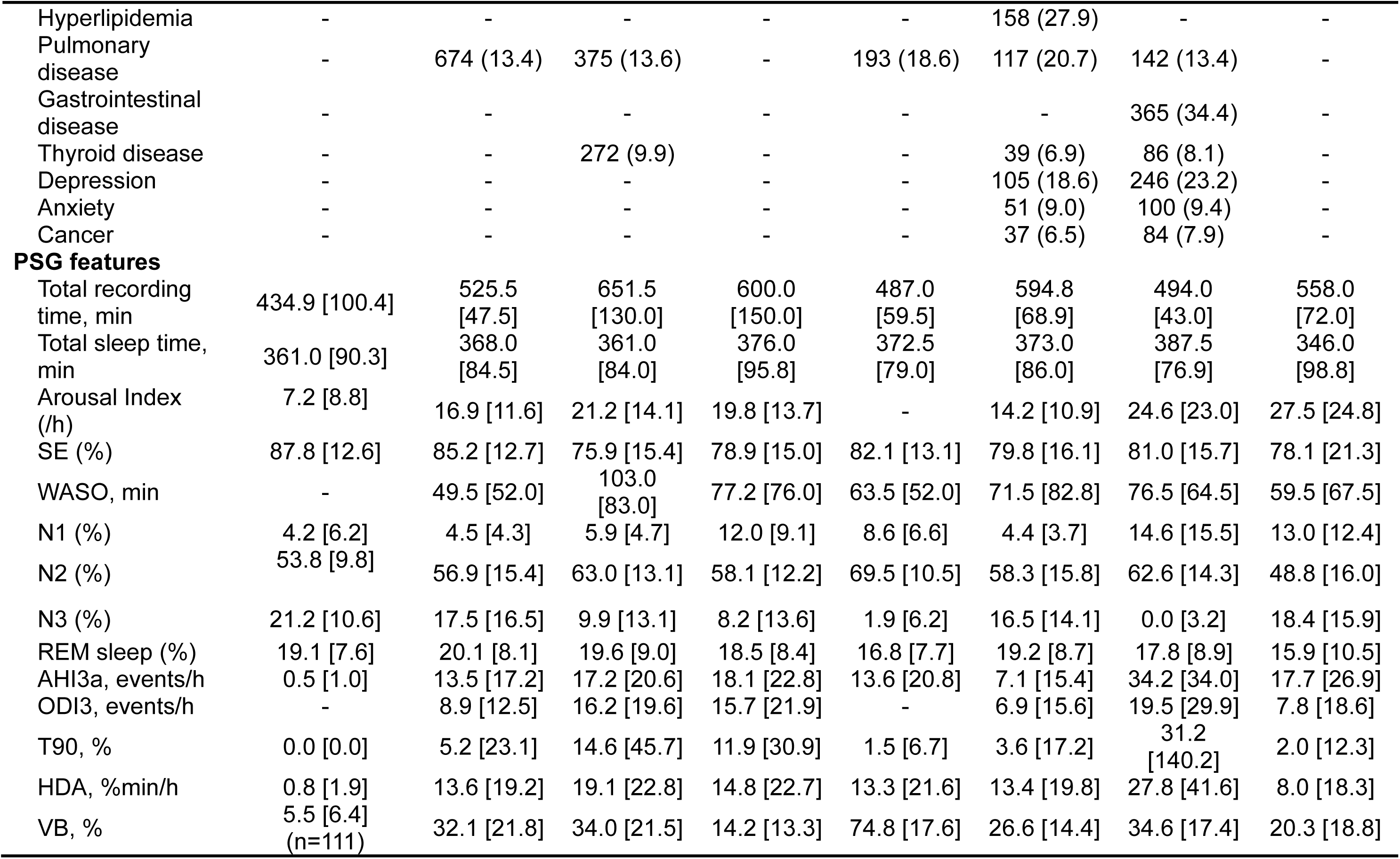
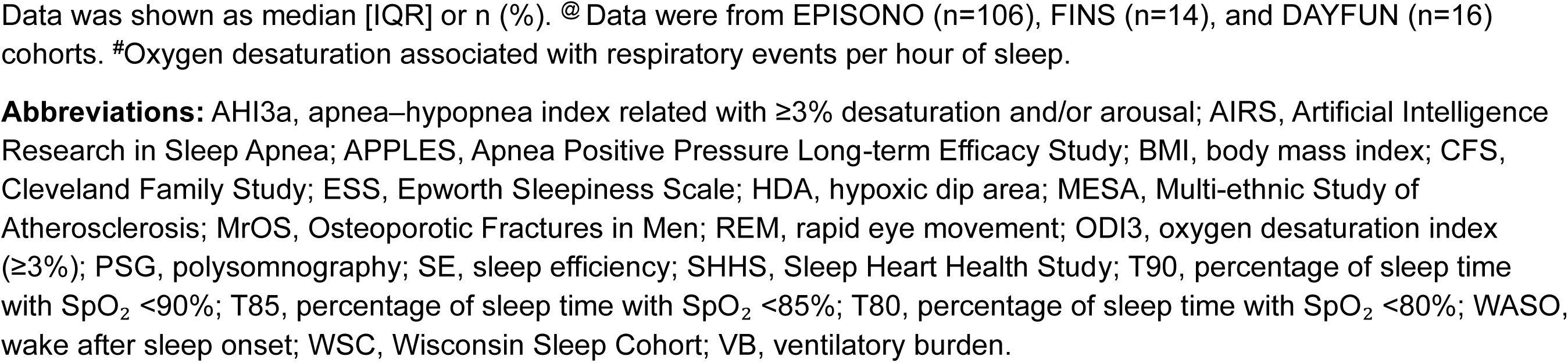
Demographic, lifestyle, and comorbidities among different datasets.

|  | Asymptomatic<br>subjects@<br>(n=136) | SHHS<br>(n=5,018) | MrOS<br>(n=2,759) | MESA<br>(n=1,326) | WSC<br>(n=1,037) | CFS<br>(n=566) | APPLES<br>(n=1,062) | AIRS<br>(n=2,263) |
| --- | --- | --- | --- | --- | --- | --- | --- | --- |
| **Demographics** |  |  |  |  |  |  |  |  |
| Age | 29.0 [14.2]<br>(20.0-52.0) | 63.0 [17.0]<br>(39.0-90.0) | 76.0 [8.0]<br>(67.0-96.0) | 67.5 [13.8]<br>(54.0-94.0) | 56.0 [12.0]<br>(37.0-82.0) | 47.0 [23.0]<br>(18.0-88.5) | 52.0 [17.0]<br>(18.0-83.0) | 54.0 [21.0]<br>(18.0-90.0) |
| Male, n (%) | 46 (33.8) | 2440<br>(48.6) | 2759<br>(100.0) | 597 (45.0) | 564 (54.4) | 249 (44.0) | 696 (65.5) | 1130 (49.9) |
| BMI | 22.9 [4.6]<br>(n=132) | 27.5 [6.0] | 26.7 [4.7] | 27.7 [7.1] | 30.1 [8.8] | 32.5 [11.7] | 30.8 [9.0] | - |
| Race | - |  |  |  |  |  |  |  |
| White | - | 4300<br>(85.7) | 2511<br>(91.0) | 478 (36.0) | 992 (95.7) | 247 (43.6) | 809 (76.2) | 839 (37.1) |
| Black | - | 388 (7.7) | 91 (3.3) | 350 (26.4) | 14 (1.4) | 306 (54.1) | 95 (8.9) | 417 (18.4) |
| Other | - | 330 (6.6) | 157 (5.7) | 498 (37.6) | 31 (3.0) | 13 (2.3) | 158 (14.9) | 1007 (44.5) |
| Smoking | n=30 |  |  |  |  |  |  |  |
| Smoker | 0 (0) | 2640<br>(52.6) | 1653<br>(59.9) | 586 (44.2) | 529 (51.0) | 150 (26.5) | 138 (13.0) | 170 (7.5) |
| Non-smoker | 30 (100) | 2349<br>(46.8) | 1106<br>(40.1) | 730 (55.1) | 508 (49.0) | 277 (48.9) | 919 (86.5) | 621 (27.4) |
| ESS total score | 6.0 [7.0]<br>(n=127) | 7.0 [7.0] | 6.0 [5.0] | 5.0 [5.0] | 9.0 [6.0] | 8.0 [7.0] | 10.0 [6.0] | 8.0 [8.0] |
| **Comorbidities, n (%)** |  |  |  |  |  |  |  |  |
| Cardiovascular disease | - | 815 (16.2) | 820 (29.7) | - | 99 (9.5) | 123 (21.7) | 421 (39.6) | - |
| Cerebrovascular disease | - | 169 (3.4) | 298 (10.8) | - | - | 16 (2.8) | 5 (0.5) | - |
| Hypertension | - | 1777<br>(35.4) | 1376<br>(49.9) | - | 339 (32.7) | 200 (35.3) | - | - |
| Diabetes mellitus | - | 338 (6.7) | 372 (13.5) | - | 115 (11.1) | 104 (18.4) | 77 (7.3) | - |
| Hyperlipidemia | - | - | - | - | - | 158 (27.9) | - | - |
| Pulmonary disease | - | 674 (13.4) | 375 (13.6) | - | 193 (18.6) | 117 (20.7) | 142 (13.4) | - |
| Gastrointestinal disease | - | - | - | - | - | - | 365 (34.4) | - |
| Thyroid disease | - | - | 272 (9.9) | - | - | 39 (6.9) | 86 (8.1) | - |
| Depression | - | - | - | - | - | 105 (18.6) | 246 (23.2) | - |
| Anxiety | - | - | - | - | - | 51 (9.0) | 100 (9.4) | - |
| Cancer | - | - | - | - | - | 37 (6.5) | 84 (7.9) | - |
| **PSG features** |  |  |  |  |  |  |  |  |
| Total recording time, min | 434.9 [100.4] | 525.5 [47.5] | 651.5 [130.0] | 600.0 [150.0] | 487.0 [59.5] | 594.8 [68.9] | 494.0 [43.0] | 558.0 [72.0] |
| Total sleep time, min | 361.0 [90.3] | 368.0 [84.5] | 361.0 [84.0] | 376.0 [95.8] | 372.5 [79.0] | 373.0 [86.0] | 387.5 [76.9] | 346.0 [98.8] |
| Arousal Index (/h) | 7.2 [8.8] | 16.9 [11.6] | 21.2 [14.1] | 19.8 [13.7] | - | 14.2 [10.9] | 24.6 [23.0] | 27.5 [24.8] |
| SE (%) | 87.8 [12.6] | 85.2 [12.7] | 75.9 [15.4] | 78.9 [15.0] | 82.1 [13.1] | 79.8 [16.1] | 81.0 [15.7] | 78.1 [21.3] |
| WASO, min | - | 49.5 [52.0] | 103.0 [83.0] | 77.2 [76.0] | 63.5 [52.0] | 71.5 [82.8] | 76.5 [64.5] | 59.5 [67.5] |
| N1 (%) | 4.2 [6.2] | 4.5 [4.3] | 5.9 [4.7] | 12.0 [9.1] | 8.6 [6.6] | 4.4 [3.7] | 14.6 [15.5] | 13.0 [12.4] |
| N2 (%) | 53.8 [9.8] | 56.9 [15.4] | 63.0 [13.1] | 58.1 [12.2] | 69.5 [10.5] | 58.3 [15.8] | 62.6 [14.3] | 48.8 [16.0] |
| N3 (%) | 21.2 [10.6] | 17.5 [16.5] | 9.9 [13.1] | 8.2 [13.6] | 1.9 [6.2] | 16.5 [14.1] | 0.0 [3.2] | 18.4 [15.9] |
| REM sleep (%) | 19.1 [7.6] | 20.1 [8.1] | 19.6 [9.0] | 18.5 [8.4] | 16.8 [7.7] | 19.2 [8.7] | 17.8 [8.9] | 15.9 [10.5] |
| AHI3a, events/h | 0.5 [1.0] | 13.5 [17.2] | 17.2 [20.6] | 18.1 [22.8] | 13.6 [20.8] | 7.1 [15.4] | 34.2 [34.0] | 17.7 [26.9] |
| ODI3, events/h | - | 8.9 [12.5] | 16.2 [19.6] | 15.7 [21.9] | - | 6.9 [15.6] | 19.5 [29.9] | 7.8 [18.6] |
| T90, % | 0.0 [0.0] | 5.2 [23.1] | 14.6 [45.7] | 11.9 [30.9] | 1.5 [6.7] | 3.6 [17.2] | 31.2 [140.2] | 2.0 [12.3] |
| HDA, %min/h | 0.8 [1.9] | 13.6 [19.2] | 19.1 [22.8] | 14.8 [22.7] | 13.3 [21.6] | 13.4 [19.8] | 27.8 [41.6] | 8.0 [18.3] |
| VB, % | 5.5 [6.4]<br>(n=111) | 32.1 [21.8] | 34.0 [21.5] | 14.2 [13.3] | 74.8 [17.6] | 26.6 [14.4] | 34.6 [17.4] | 20.3 [18.8] |

Data was shown as median [IQR] or n (%). @ Data were from EPISONO (n=106), FINS (n=14), and DAYFUN (n=16) cohorts. #Oxygen desaturation associated with respiratory events per hour of sleep.
**Abbreviations:** AHI3a, apnea–hypopnea index related with $\geq 3\%$ desaturation and/or arousal; AIRS, Artificial Intelligence Research in Sleep Apnea; APPLES, Apnea Positive Pressure Long-term Efficacy Study; BMI, body mass index; CFS, Cleveland Family Study; ESS, Epworth Sleepiness Scale; HDA, hypoxic dip area; MESA, Multi-ethnic Study of Atherosclerosis; MrOS, Osteoporotic Fractures in Men; REM, rapid eye movement; ODI3, oxygen desaturation index ( $\geq 3\%$ ); PSG, polysomnography; SE, sleep efficiency; SHHS, Sleep Heart Health Study; T90, percentage of sleep time with $\text{SpO}_2 < 90\%$ ; T85, percentage of sleep time with $\text{SpO}_2 < 85\%$ ; T80, percentage of sleep time with $\text{SpO}_2 < 80\%$ ; WASO, wake after sleep onset; WSC, Wisconsin Sleep Cohort; VB, ventilatory burden.

### Normative reference value of HDA

All 136 observations were used to derive the HDA cutoff. Figure 1 upper panel shows the distribution of HDA among asymptomatic individuals, demonstrating a right-skewed distribution. The red dashed line indicates the 97.5^th^ percentile cutoff of HDA (8.60%min/h). The lower panel illustrates the distribution of HDA across the combined cohorts with 14,031 subjects. The distribution is markedly right-skewed, with a wide range of values. The mean HDA was 23.6 (SD: 30.7), the median was 14.7 (IQR: 22.3), and the values ranged from 0 to 502.4. This cutoff of 8.6%min/h was then applied to combined cohort to classify subjects into 2 groups: 4,500 subjects with normal HDA and 9,531 subjects with elevated HDA (Supplementary Table 1).

**Figure 1.**
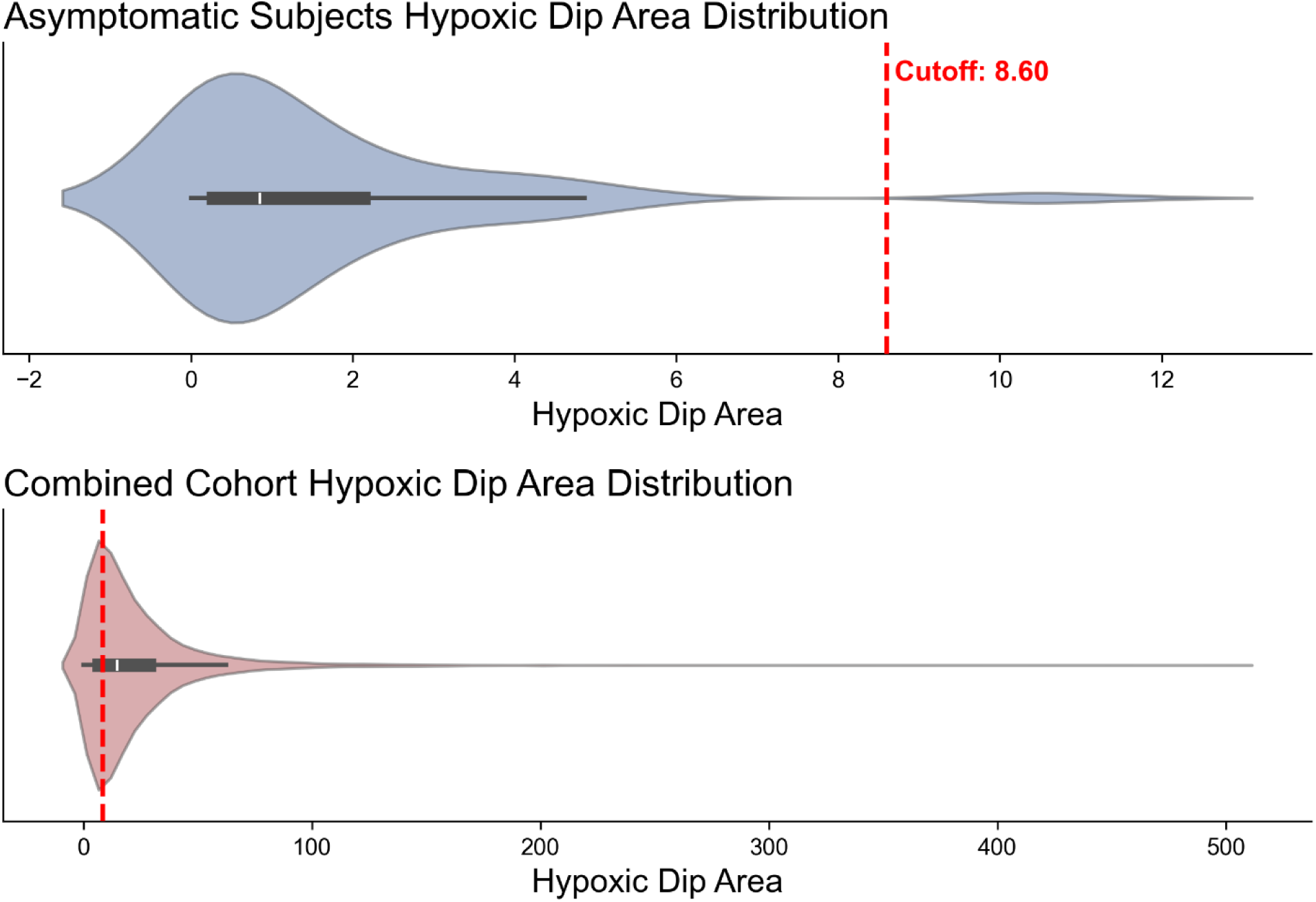
Distribution of hypoxic dip area among asymptomatic individuals and the combined cohorts, including SHHS, MrOS, MESA, WSC, CFS, APPLES, and AIRS. **Abbreviations:** AIRS, Artificial Intelligence Research in Sleep Apnea; APPLES, Apnea Positive Pressure Long-term Efficacy Study; CFS, Cleveland Family Study; MESA, Multi-ethnic Study of Atherosclerosis; MrOS, Osteoporotic Fractures in Men; SHHS, Sleep Heart Health Study; WSC, Wisconsin Sleep Cohort.

Individuals with elevated HDA were older and more frequently male compared with those with normal HDA (median age 67.0 vs 58.0 years; 65.9% vs 47.9%), and had higher BMI (supplementary table 1). Racial distribution and smoking status also differed between groups, with a higher proportion of White subjects and smokers in the elevated HDA group. In terms of sleep characteristics, subjects with elevated HDA exhibited poorer sleep quality, including shorter total sleep time (362.5 [IQR: 87.8] vs 373.5 [IQR: 87.0] mins), lower sleep efficiency (79.8% [IQR: 15.9] vs 83.4% [IQR: 15.1]), and greater wake after sleep onset (72.5 [IQR: 71.5] vs 53.0 [IQR: 60.0] mins). Sleep architecture was altered, with increased proportions of lighter sleep stages (N1 and N2) and reduced deep sleep (N3) and REM sleep. Both HDA and VB were markedly higher in the elevated HDA group (HDA: 22.4 [IQR: 22.9] vs 4.3 [IQR: 4.0] %min/h; VB: 33.2% [IQR: 23.5] vs 22.9% [IQR: 23.8]), indicating more severe nocturnal hypoxemia and ventilatory instability.

### Respiration, arousal, and oxygenation profiles between normal and elevated HDA

Compared with subjects with normal HDA, those with elevated HDA exhibited markedly worse respiratory and oxygenation profiles (Table 2). The elevated HDA group had significantly higher AHI3a (22.2 [IQR: 20.2] vs 6.8 [IQR: 10.2] events/h) and arousal index (21.5 [IQR: 16.3] vs 16.5 [IQR: 12.3] events/h; both P<0.001). Oxygenation metrics were consistently impaired, with lower mean SpO_2_ (93.4% [IQR: 2.2] vs 95.2% [IQR: 1.7]) and markedly greater nocturnal hypoxemia, as reflected by higher T90, T85, and T80 (all P<0.001). Similarly, ODI3 was significantly higher in the elevated HDA group (17.0 [IQR: 19.8] vs 3.7 [IQR: 4.8] events/h, P<0.001). These differences remained highly significant after adjustment for additional covariates. To assess whether these findings were consistent across cohorts and not driven by any single cohort, we performed cohort-specific analyses. As shown in Figure 2, the observed patterns were consistent across cohorts.

**Figure 2.**
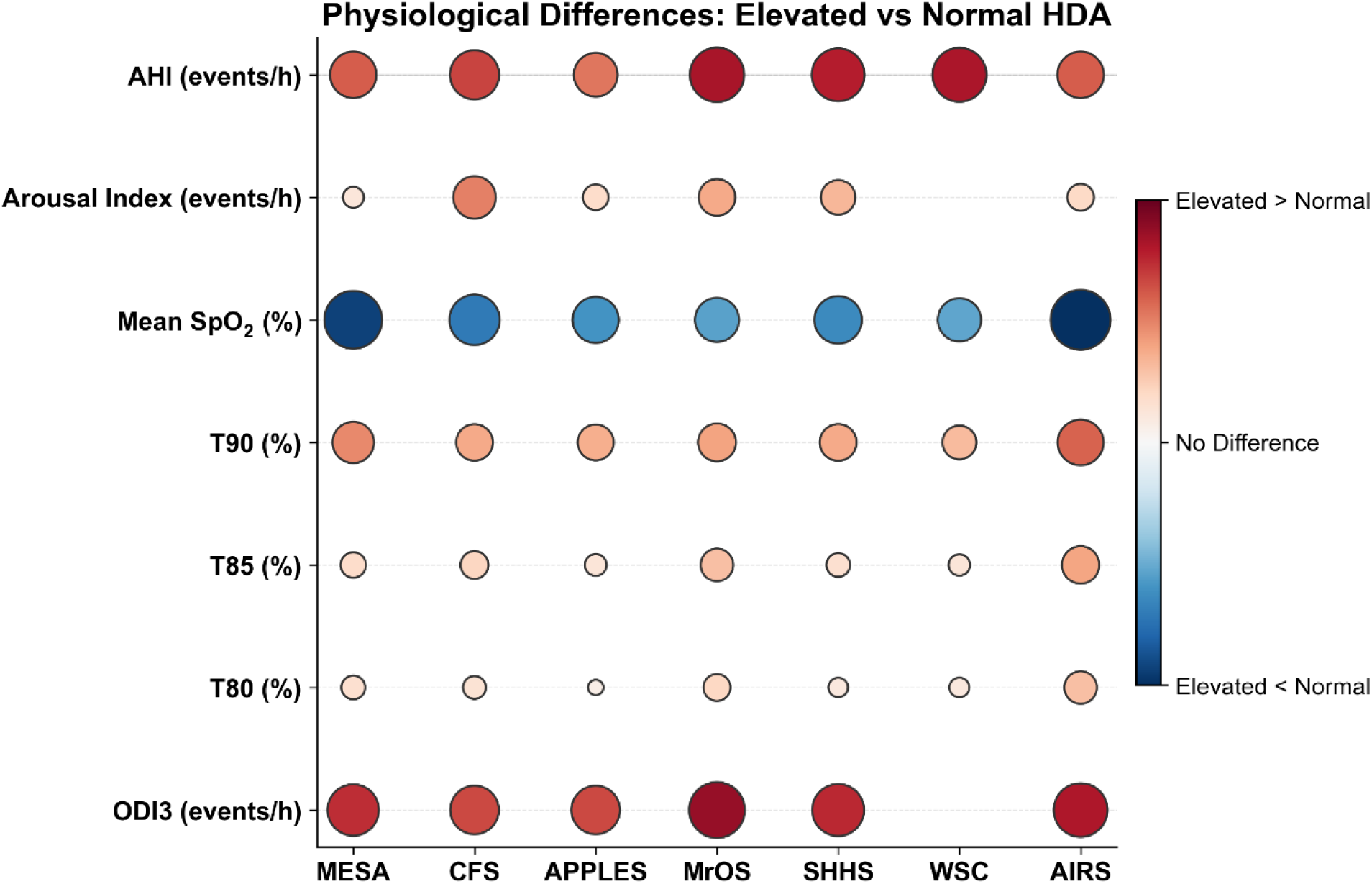
Cohort-specific analyses demonstrating consistent differences in respiratory, arousal, and oxygenation profiles between subjects with normal and elevated HDA across cohorts. The models were separately generated by linear regression models with adjustment of age, sex, body mass index, race, smoking status, total recording time, and ventilatory burden. **Abbreviations:** AHI3a, apnea–hypopnea index related with ≥3% desaturation and/or arousal; AIRS, Artificial Intelligence Research in Sleep Apnea; APPLES, Apnea Positive Pressure Long-term Efficacy Study; CFS, Cleveland Family Study; HDA, hypoxic dip area; MESA, Multi-ethnic Study of Atherosclerosis; MrOS, Osteoporotic Fractures in Men; ODI3, oxygen desaturation index (≥3%); SHHS, Sleep Heart Health Study; T90, percentage of sleep time with SpO₂ <90%; T85, percentage of sleep time with SpO₂ <85%; T80, percentage of sleep time with SpO₂ <80%; WSC, Wisconsin Sleep Cohort.

**Table 2.**
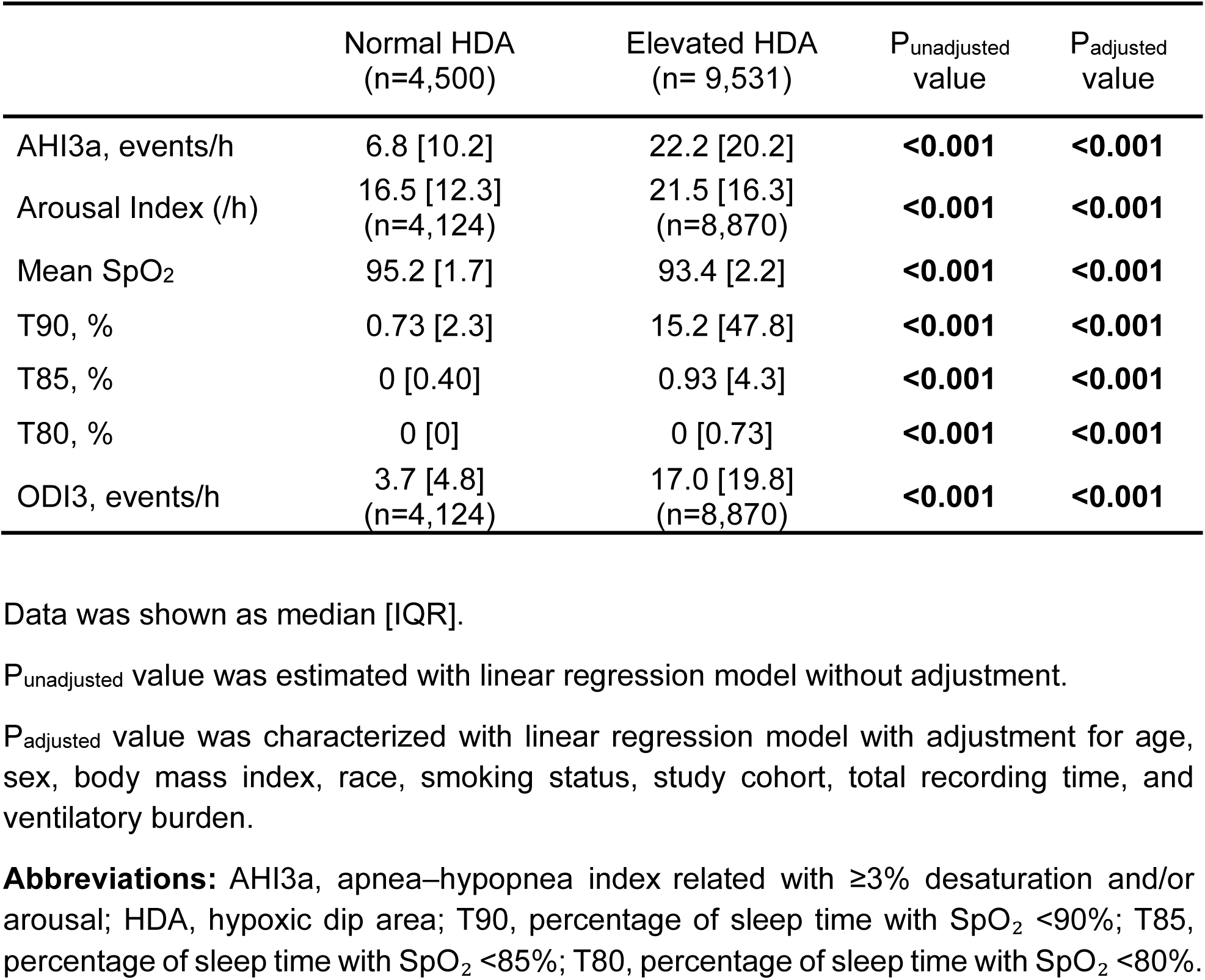
Respiratory, arousal, and oxygenation profiles between normal and elevated HDA subjects.

|  | Normal HDA<br>(n=4,500) | Elevated HDA<br>(n= 9,531) | P<sub>unadjusted</sub><br>value | P<sub>adjusted</sub><br>value |
| --- | --- | --- | --- | --- |
| AHI3a, events/h | 6.8 [10.2] | 22.2 [20.2] | **<0.001** | **<0.001** |
| Arousal Index (/h) | 16.5 [12.3]<br>(n=4,124) | 21.5 [16.3]<br>(n=8,870) | **<0.001** | **<0.001** |
| Mean SpO<sub>2</sub> | 95.2 [1.7] | 93.4 [2.2] | **<0.001** | **<0.001** |
| T90, % | 0.73 [2.3] | 15.2 [47.8] | **<0.001** | **<0.001** |
| T85, % | 0 [0.40] | 0.93 [4.3] | **<0.001** | **<0.001** |
| T80, % | 0 [0] | 0 [0.73] | **<0.001** | **<0.001** |
| ODI3, events/h | 3.7 [4.8]<br>(n=4,124) | 17.0 [19.8]<br>(n=8,870) | **<0.001** | **<0.001** |

Data was shown as median [IQR].
P<sub>unadjusted</sub> value was estimated with linear regression model without adjustment.
P<sub>adjusted</sub> value was characterized with linear regression model with adjustment for age, sex, body mass index, race, smoking status, study cohort, total recording time, and ventilatory burden.
**Abbreviations:** AHI3a, apnea–hypopnea index related with $\geq 3\%$ desaturation and/or arousal; HDA, hypoxic dip area; T90, percentage of sleep time with SpO<sub>2</sub> <90%; T85, percentage of sleep time with SpO<sub>2</sub> <85%; T80, percentage of sleep time with SpO<sub>2</sub> <80%.

To assess the robustness of our findings, we performed sensitivity analyses using age-, sex-, and BMI-adjusted threshold derived from multivariable regression model. The results were consistent, showing similar relationships when the adjusted threshold was applied (Supplementary Table 2).

### EDS, clinical comorbidities, and incident events between normal and elevated HDA

Compared with subjects with normal HDA, those with elevated HDA had a marginally higher percentage of EDS (32.6% vs 31.0%, P_adjusted_ =0.001) (Table 3). Pre-existing CVD and hypertension were significantly more prevalent in the elevated HDA group (57.9% vs 35.1% and 40.9% vs 26.6%, respectively), and these patterns remained significant after adjustment for covariates. Other conditions, including cerebrovascular disease, diabetes mellitus, hyperlipidemia, pulmonary disease, thyroid disease, and cancer, were not significantly different after adjustment. Sensitivity analyses using age-, sex-, and BMI-adjusted threshold derived from multivariable regression models showed similar results (Supplementary Table 3).

**Table 3.** Excessive daytime sleepiness, clinical comorbidities and incident events between normal and elevated HDA subjects.

|  | Normal HDA<br>(n=4,500) | Elevated HDA<br>(n=9,531) | P<sub>unadjusted</sub><br>value | P<sub>adjusted</sub><br>value |
| --- | --- | --- | --- | --- |
| Excessive daytime sleepiness (ESS ≥ 10) | 1394 (31.0) | 3106 (32.6) | 0.056 | **0.001** |
| **Clinical diseases, lifetime** |  |  |  |  |
| Cardiovascular disease | 278 (35.1)<br>(n=792) | 1180 (57.9)<br>(n=2,039) | **<0.001** | **0.006** |
| Cerebrovascular disease | 37 (5.6)<br>(n=661) | 153 (8.1)<br>(n=1,890) | **0.035** | 0.95 |
| Hypertension | 582 (26.6)<br>(n=2,187) | 1734 (40.9)<br>(n=4,244) | **<0.001** | **0.004** |
| Diabetes mellitus | 128 (5.5)<br>(n=2,323) | 506 (9.8)<br>(n=5,149) | **<0.001** | 0.85 |
| Hyperlipidemia | 33 (17.3)<br>(n=191) | 125 (34.2)<br>(n=366) | **<0.001** | 0.52 |
| Pulmonary disease | 316 (13.4)<br>(n=2,350) | 810 (15.6)<br>(n=5,176) | **0.013** | 0.054 |
| Gastrointestinal disease | 46 (30.9)<br>(n=149) | 319 (35.4)<br>(n=902) | 0.29 | 0.66 |
| Thyroid disease | 17 (5.0)<br>(n=342) | 108 (8.5)<br>(n=1,271) | **0.032** | 0.19 |
| Depression | 77 (22.6)<br>(n=340) | 274 (21.5)<br>(n=1,272) | 0.66 | 0.79 |
| Anxiety | 40 (11.8)<br>(n=339) | 111 (8.8)<br>(n=1,265) | 0.092 | 0.24 |
| Cancer | 14 (4.1)<br>(n=342) | 107 (8.4)<br>(n=1,272) | **0.008** | 0.37 |
| **Incident event** |  |  |  |  |
| Cardiovascular disease | 344 (10.2)<br>(n = 3,358) | 1,287 (19.7)<br>(n = 6,541) | - | - |
| All-cause mortality | 620 (29.0)<br>(n = 2,139) | 2,441 (46.3)<br>(n = 5,269) | - | - |
| Cardiovascular disease mortality | 185 (11.0)<br>(n = 1,686) | 819 (21.6)<br>(n = 3,841) | - | - |

Data was shown as n (%).
P<sub>unadjusted</sub> value was calculated with logistic regression model without adjustment.
P<sub>adjusted</sub> value was measured with logistic regression model with adjustment for age, sex, body mass index, race, smoking status, study cohort, total recording time, and ventilatory burden.
**Abbreviations:** ESS, Epworth Sleepiness Scale; HDA, hypoxic dip area.

For incident outcomes, subjects with elevated HDA had significantly higher proportions of CVD events (19.7% vs 10.2%), all-cause mortality (46.3% vs 29.0%), and CVD mortality (21.6% vs 11.0%) (Table 3). In Cox proportional hazards analyses, subjects with elevated HDA had significantly higher risks of incident CVD disease (HR, 1.86; 95% CI, 1.66-2.10), all-cause mortality (HR, 1.61; 95% CI, 1.48-1.76), and CVD mortality (HR, 1.91; 95% CI, 1.63-2.25) in unadjusted models (all P<0.001). However, these associations were attenuated and were no longer statistically significant after adjustment for covariates (Table 4). Cohort-specific analyses showed similar findings, with elevated HDA associated with higher risks of incident events in unadjusted analyses. These associations were generally attenuated after multivariable adjustment, although the association with incident CVD remained significant in the MrOS cohort (Supplementary Table 4).

**Table 4.** Incident events between normal and elevated HDA subjects.

|  | Unadjusted model |  | Adjusted model |  |
| --- | --- | --- | --- | --- |
|  | HR (95% CI) | P value | HR (95% CI) | P value |
| Cardiovascular disease | 1.86 (1.66, 2.10) | **<0.001** | 1.09 (0.95, 1.24) | 0.21 |
| All-cause mortality | 1.61 (1.48, 1.76) | **<0.001** | 1.03 (0.94, 1.13) | 0.47 |
| Cardiovascular disease mortality | 1.91 (1.63, 2.25) | **<0.001** | 1.09 (0.92, 1.28) | 0.30 |

P<sub>unadjusted</sub> value was calculated with Cox regression model without adjustment.
P<sub>adjusted</sub> value was measured with Cox regression model with adjustment for age, sex, body mass index, race, smoking status, study cohort, total recording time, and ventilatory burden.
**Abbreviations:** HDA, hypoxic dip area.

## Discussion

Area-based hypoxic burden is emerging as a superior alternative to AHI for OSA severity assessment. Yet, a critical translational gap remains: without a physiologically meaningful threshold, the clinical utility of hypoxic burden cannot move from associational findings to actionable, real-world implementation in clinical management of OSA. To help address this gap, we adapted a well-established principle from other fields of medicine, namely the derivation of normative reference values, and applied it to sleep medicine. Using a fully automated area-based hypoxic burden (i.e. HDA) derived from 136 asymptomatic individuals representing a physiologically normal, OSA-free population, we established reference boundaries distinguishing normal from elevated hypoxic burden. Strikingly, a cutoff of 8.6 %min/h robustly stratified individuals across respiratory, arousal, and hypoxemia-related physiological dimensions. This threshold demonstrated consistent discriminative performance across 7 independent cohorts, underscoring its generalizability and potential as a clinically deployable benchmark.

HDA differs from conventional OSA metrics by integrating both the depth and duration of desaturation events through quantification of the area under the oxygen desaturation curve.^7,10,15,16^ Unlike the AHI and ODI, which primarily reflect event frequency, or T90/T85/T80, which measure cumulative time below fixed saturation thresholds, HDA captures the full morphology of desaturation and reoxygenation events. This integrated characterization may explain its strong connection with physiological severity across respiratory, arousal, and oxygenation domains observed in this study.

Our study showed that although elevated HDA, defined by a physiologically derived cutoff, was strongly allied with more severe nocturnal hypoxemia, increased respiratory events, arousal, and daytime sleepiness, and was related with increased prevalence of incident CVD outcomes, upon adjusting for known covariates, the difference was no longer statistically significant. These findings highlight a critical distinction between physiologically derived threshold for routine clinical management and outcome-driven risk threshold. Prior studies have supported that risk signals are primarily observed in individuals within the highest HDA categories (e.g., top quintile), corresponding to significantly higher thresholds (e.g., 71%min/h).^10,17^ In contrast, the cutoff of 8.6%min/h derived from asymptomatic individuals in our study represents a lower, physiology-based threshold that effectively delineates abnormal/elevated HDA, but does not confer prognostic discrimination. These findings might support a two-threshold framework, in which physiologically derived cutoff is used to define disease severity, characterize phenotypes, and inform disease diagnosis and monitoring, whereas separate, outcome-driven threshold may be required for risk stratification of long-term clinical outcomes (Figure 3). Similar distinctions have been recognized in other diseases; for example, hemoglobin A1c in diabetes mellitus uses a threshold of ≥6.5% for diagnosis, whereas lower thresholds (e.g., ≥5.7%) define prediabetes and are used to stratify future cardiovascular and metabolic risk.^44,45^

**Figure 3.**
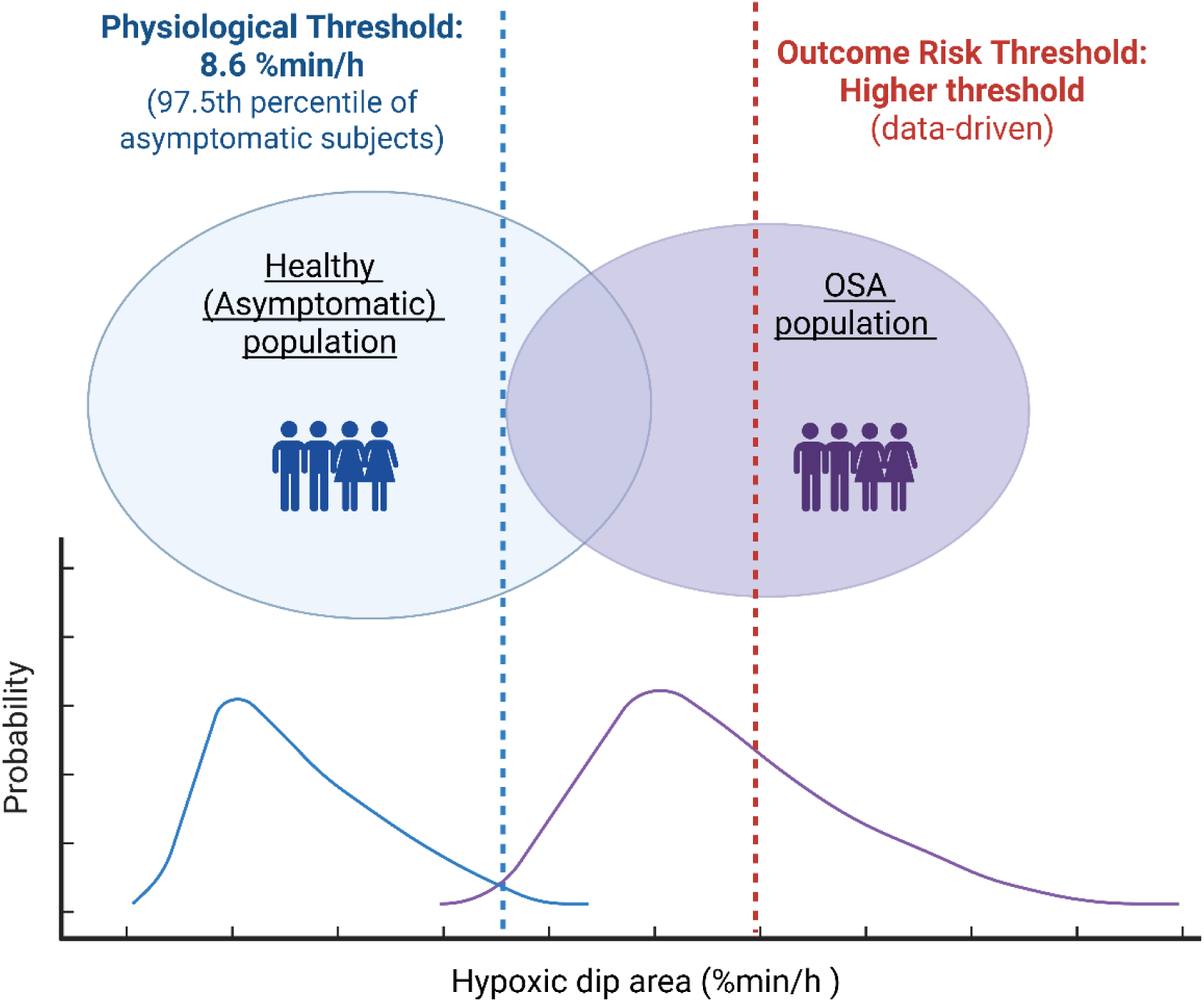
Schematic illustration of the proposed two-threshold concept for hypoxic dip area. **Abbreviations:** OSA, obstructive sleep apnea.

Our study has several strengths. First, we leveraged multiple large community-based and clinical cohorts, enhancing the generalizability of our findings. Second, we derived normative reference values from asymptomatic individuals without evidence of OSA and other respiratory diseases, providing a physiologically grounded approach to defining HDA physiological cutoff. Third, the robustness of our findings was supported by sensitivity analyses. Several limitations should be noted. First, although we included a modest sample of asymptomatic healthy individuals, those aged >55 years were underrepresented, which may limit generalizability across the full age spectrum. Second, treatment effects were not accounted for in the analysis of mortality outcomes. Third, cognitive outcomes were not evaluated. Future studies incorporating cognitive function are warranted to further validate the proposed cutoff.

## Conclusions

Our study derived a physiological cutoff of 8.6%min/h for HDA from asymptomatic individuals that reliably distinguishes physiological severity across respiratory, arousal, and oxygenation domains and generalizes across multiple cohorts. This physiologically derived threshold did not independently predict incident cardiovascular events or mortality. These findings support the use of HDA for phenotyping physiological severity of hypoxemia, while highlighting the need for outcome-driven thresholds to enable long-term outcome risk stratification.

## Funding

This study was funded by National Institutes of Health (R01HL171813, K25HL151912, R21HL165320, R21HL173733 R01HL081310, and R21AG059179).

## Disclosure statements

The authors have no financial arrangements or connections. The authors have no potential conflicts of interest.

## Data availability

Data from the NSRR are available upon request through the NSRR website. Data from the AIRS cohort are available from the corresponding author upon reasonable request.

## Code availability

The source code for the hypoxic dip area extraction algorithm is available at: https://github.com/SleepAndCircadianAnalysis/hypoxicburden.

## Acknowledgements

We would like to thank our research staff and all subjects for their contributions to this study. The Sao Paulo Epidemiologic Study (EPISONO) was supported by the Associação Fundo de Incentivo à Pesquisa, São Paulo (AFIP). ST is a recipient of a Conselho Nacional de Desenvolvimento Científico e Tecnológico (CNPq) fellowship. MLA is a recipient of a CNPq fellowship and a grant from the Fundação de Amparo à Pesquisa do Estado de São Paulo (FAPESP #2020/13467-8). The National Sleep Research Resource was supported by the U.S. National Institutes of Health, National Heart Lung and Blood Institute (R24 HL114473, 75N92019R002). The Sleep Heart Health Study (SHHS) was supported by National Heart, Lung, and Blood Institute cooperative agreements U01HL53916 (University of California, Davis), U01HL53931 (New York University), U01HL53934 (University of Minnesota), U01HL53937 and U01HL64360 (Johns Hopkins University), U01HL53938 (University of Arizona), U01HL53940 (University of Washington), U01HL53941 (Boston University), and U01HL63463 (Case Western Reserve University). The Osteoporotic Fractures in Men (MrOS) Study is supported by National Institutes of Health funding. The following institutes provide support: the National Institute on Aging (NIA), the National Institute of Arthritis and Musculoskeletal and Skin Diseases (NIAMS), the National Center for Advancing Translational Sciences (NCATS), and NIH Roadmap for Medical Research under the following grant numbers: U01 AG027810, U01 AG042124, U01 AG042139, U01 AG042140, U01 AG042143, U01 AG042145, U01 AG042168, U01 AR066160, R01 AG066671, and UL1 TR002369. The National Heart, Lung, and Blood Institute (NHLBI) provides funding for the MrOS Sleep ancillary study “Outcomes of Sleep Disorders in Older Men” under the following grant numbers: R01 HL071194, R01 HL070848, R01 HL070847, R01 HL070842, R01 HL070841, R01 HL070837, R01 HL070838, and R01 HL070839. The Multi-Ethnic Study of Atherosclerosis (MESA) Sleep Ancillary study was funded by NIH-NHLBI Association of Sleep Disorders with Cardiovascular Health Across Ethnic Groups (RO1 HL098433). MESA is supported by NHLBI funded contracts HHSN268201500003I, N01-HC-95159, N01-HC-95160, N01-HC-95161, N01-HC-95162, N01-HC-95163, N01-HC-95164, N01-HC-95165, N01-HC-95166, N01-HC-95167, N01-HC-95168 and N01-HC-95169 from the National Heart, Lung, and Blood Institute, and by cooperative agreements UL1-TR-000040, UL1-TR-001079, and UL1-TR-001420 funded by NCATS. The Cleveland Family Study (CFS) was supported by grants from the National Institutes of Health (HL46380, M01 RR00080-39, T32-HL07567, RO1-46380). This Wisconsin Sleep Cohort Study was supported by the U.S. National Institutes of Health, National Heart, Lung, and Blood Institute (R01HL62252), National Institute on Aging (R01AG036838, R01AG058680), and the National Center for Research Resources (1UL1RR025011). The Apnea Positive Pressure Long-term Efficacy Study (APPLES) was supported by the National Heart, Lung, and Blood Institute (U01HL68060).

## Author contributions

L.Z. and A.P. contributed to the conception and design of the study. L.Z., S.Y.Y., S. D.W., P.B., E.H., Z.R., S.C., A.K., T.M.T., K.K., A.W.V., L.B.M.D, L.O.P., M.L.A., S.T., K.L.S., I.A., D.M.R., and A.P. contributed to data collection/management. L.Z., S.Y.Y., S.D.W., P.B., and A.P. handled data cleansing, statistical analysis, and data interpretation. L.Z. and A.P. wrote the main paper. All authors have reviewed and approved the final paper.

## Notes

### Competing Interest Statement

The authors have declared no competing interest.

### Author Declarations

Ethics committee/IRB of Mount Sinai gave ethical approval for this work

