## Supplementary information for "Establishing a physiological normative value of area-based hypoxic burden in obstructive sleep apnea"

**Supplementary Table 1 Demographics and sleep features between normal and elevated HDA subjects.**

|  | Normal HDA  (n=4,500) | Elevated HDA  (n=9,531) |
| --- | --- | --- |
| Demographics |  |  |
| Age | 58.0 [22.0] | 67.0 [18.0] |
| Male, n (%) | 2,156 (47.9) | 6,279 (65.9) |
| BMI | 26.1 [5.6]  (n=3,330) | 28.6 [6.7]  (n=8,438) |
| Race |  |  |
| White | 3,002 (66.7) | 7,174 (75.3) |
| Black | 582 (12.9) | 1,079 (11.3) |
| Other | 916 (20.4) | 1,278 (13.4) |
| Smoking |  |  |
| Smoker | 1,589 (43.2) | 4,277 (49.2) |
| Non-smoker | 2,093 (56.8) | 4,417 (50.8) |
| PSG sleep features |  |  |
| Total recording time, min | 534.5 [109.0] | 534.5 [110.5] |
| Total sleep time, min | 373.5 [87.0] | 362.5 [87.8] |
| SE (%) | 83.4 [15.1] | 79.8 [15.9] |
| WASO, min | 53.0 [60.0] | 72.5 [71.5] |
| N1 (%) | 6.3 [7.2] | 7.3 [8.2] |
| N2 (%) | 56.7 [16.2] | 59.8 [16.4] |
| N3 (%) | 15.7 [17.4] | 10.9 [17.3] |
| REM (%) | 19.4 [8.8] | 18.4 [9.1] |
| HDA, %min/h | 4.3 [4.0] | 22.4 [22.9] |
| VB, % | 22.9 [23.8] | 33.2 [23.5] |

Data was shown as median [IQR] or n (%).

P value was estimated with Mann-Whitney U test or Chi-square test.

**Abbreviations:** BMI, body mass index; HDA, hypoxic dip area; REM, rapid eye movement; SE, sleep efficiency; VB, ventilatory burden; WASO, wake after sleep onset.

**Supplementary Table 2 Respiratory,** **arousal, and oxygenation profiles between normal and elevated HDA subjects using regression analyses.**

|  | Normal HDA  (n=3,471) | Elevated HDA  (n=8,297) | P_unadjusted_ value | P_adjusted_ value |
| --- | --- | --- | --- | --- |
| AHI3a, events/h | 6.3 [7.8] | 21.9 [23.1] | **<0.001** | **<0.001** |
| Arousal Index (/hr.) | 15.3 [10.2]  (n = 3,083) | 20.6 [14.7]  (n = 7,648) | **<0.001** | **<0.001** |
| Mean SpO_2_ | 94.9 [1.8] | 93.3 [2.2] | **<0.001** | **<0.001** |
| T90 | 1.0 [2.8] | 16.3 [50.7] | **<0.001** | **<0.001** |
| T85 | 0 [0.5] | 0.95 [4.2] | **<0.001** | **<0.001** |
| T80 | 0 [0] | 0 [0.70] | **<0.001** | **<0.001** |
| ODI3, events/h | 4.2 [4.6]  (n=3,083) | 17.0 [19.2]  (n=7,648) | **<0.001** | **<0.001** |

Data was shown as median [IQR].

P_unadjusted_ value was estimated with linear regression model without adjustment.

P_adjusted_ value was calculated with linear regression model with adjustment for age, sex, body mass index, race, smoking status, study cohort, total recording time, and ventilatory burden.

**Abbreviations:** AHI3a, apnea–hypopnea index related with ≥3% desaturation and/or arousal; HDA, hypoxic dip area; ODI3, oxygen desaturation index (≥3%); SE, sleep efficiency; T90, percentage of sleep time with SpO₂ <90%; T85, percentage of sleep time with SpO₂ <85%; T80, percentage of sleep time with SpO₂ <80%; WASO, wake after sleep onset.

**Supplementary Table 3 History of clinical diseases and incident events between normal and elevated HDA subjects using regression analyses.**

|  | Normal HDA  (n=3,471) | Elevated HDA  (n=8,297) | P_unadjusted_ value | P_adjusted_ value |
| --- | --- | --- | --- | --- |
| Excessive daytime sleepiness | 949 (27.3) | 2614 (31.5) | **<0.001** | **<0.001** |
| **Clinical diseases, lifetime** |  |  |  |  |
| Cardiovascular disease | 290 (35.5)  (n=818) | 1,168 (58.0)  (n=2,013) | **<0.001** | **0.004** |
| Cerebrovascular disease | 40 (5.8)  (n=691) | 150 (8.1)  (n=1,860) | 0.053 | 0.94 |
| Hypertension | 605 (26.7)  (n=2,268) | 1,711 (41.1)  (n=4,163) | **<0.001** | **0.002** |
| Diabetes Mellitus | 129 (5.3)  (n=2,412) | 505 (10.0)  (n=5,060) | **<0.001** | 0.75 |
| Hyperlipidemia | 32 (16.5)  (n=194) | 126 (34.7)  (n=363) | **<0.001** | 0.26 |
| Pulmonary disease | 328 (13.4)  (n=2,439) | 798 (15.7)  (n=5,087) | **0.011** | 0.065 |
| Gastrointestinal disease | 47 (29.7)  (n=158) | 318 (35.6)  (n=902) | 0.15 | 0.43 |
| Thyroid disease | 17 (4.8)  (n=354) | 108 (8.6)  (n=1,259) | **0.021** | 0.14 |
| Depression | 78 (22.4)  (n=352) | 272 (21.6)  (n=1,260) | 0.73 | 0.79 |
| Anxiety | 40 (11.4)  (n=351) | 111 (8.9)  (n=1,253) | 0.15 | 0.34 |
| Cancer | 17 (4.8)  (n=354) | 104 (8.3)  (n=1,260) | **0.031** | 0.78 |
| **Incident event** |  |  |  |  |
| Cardiovascular disease | 340 (14.8)  (n=2,297) | 1,202 (22.5)  (n=5,339) | - | - |
| All-cause mortality | 663 (29.6)  (n=2,243) | 2,398 (46.4)  (n=5,165) | - | - |
| Cardiovascular disease mortality | 199 (11.3)  (n=1,764) | 805 (21.4)  (n=3,763) | - | - |

Data was shown as n (%).

P_unadjusted_ value was estimated with logistic regression model without adjustment.

P_adjusted_ value was estimated with logistic regression model with adjustment for age, sex, body mass index, race, smoking status, study cohort, total recording time, and ventilatory burden.

**Abbreviations:** HDA, hypoxic dip area.

**Supplementary Table 4** Incident events between normal and elevated HDA subjects among different cohorts.

|  | SHHS | | | | |  | MrOS | | | | |  | AIRS | | | | |
| --- | --- | --- | --- | --- | --- | --- | --- | --- | --- | --- | --- | --- | --- | --- | --- | --- | --- |
|  | Unadjusted model | |  | Adjusted model | |  | Unadjusted model | |  | Adjusted model | |  | Unadjusted model | |  | Adjusted model | |
|  | HR (95% CI) | P value |  | HR (95% CI) | P value |  | HR (95% CI) | P value |  | HR (95% CI) | P value |  | HR (95% CI) | P value |  | HR (95% CI) | P value |
| CVD | 1.71 (1.49, 1.98) | **<0.001** |  | 0.96 (0.82, 1.12) | 0.59 |  | 1.55 (1.21, 1.99) | **<0.001** |  | 1.45 (1.12, 1.87) | **0.004** |  | 3.22 (1.99, 5.21) | **<0.001** |  | 1.68 (0.70, 4.04) | 0.25 |
| All-cause mortality | 1.75 (1.52, 2.01) | **<0.001** |  | 1.01 (0.87, 1.17) | 0.94 |  | 1.14 (1.02, 1.28) | **0.023** |  | 1.06 (0.94, 1.19) | 0.35 |  | - | - |  | - | - |
| CVD mortality | 1.89 (1.43, 2.49) | **<0.001** |  | 0.99 (0.74, 1.33) | 0.96 |  | 1.31 (1.08, 1.60) | **0.007** |  | 1.15 (0.94, 1.40) | 0.17 |  | - | - |  | - | - |

P_unadjusted_ value was calculated with Cox regression model without adjustment.

P_adjusted_ value was measured with Cox regression model with adjustment for age, sex, body mass index, race, smoking status, study cohort, total recording time, and ventilatory burden.

**Abbreviations:** AIRS, Artificial Intelligence Research in Sleep Apnea; CVD, cardiovascular disease; HDA, hypoxic dip area;

MrOS, Osteoporotic Fractures in Men; SHHS, Sleep Heart Health Study.

**Supplementary methods**

**Sau Paulo Epidemiologic Sleep Study (EPISONO)**

The EPISONO study was an epidemiological cohort study conducted in 2007 to characterize the epidemiologic profile of sleep disorders in the adult population of São Paulo, Brazil. Methodological details of the EPISONO study have been described previously.^1^ This study was approved by the Ethics Committee for Research of the Universidade Federal de Sao Paulo at the Hospital Sao Paulo (CEP 0593/06). For the present study, 106 asymptomatic normal subjects were selected from the original cohort of 1,042 participants and included in the final analyses. The selection criteria and methodology used to define this asymptomatic subgroup have also been described previously.^2^ All subjects underwent attended nocturnal polysomnography (NPSG) using the EMBLA S7000 system (Embla Systems Inc., Broomfield, CO, USA).

**Relating Sleep Disordered Breathing to Daytime Function (DAYFUN)**

The DAYFUN study involved subjects evaluated for OSA at the New York University (NYU) Sleep Disorders Center as well as healthy community-dwelling adults aged >18 years. Healthy subjects had no evidence of sleep disorders, witnessed apneas, or snoring based on sleep physician clinical interviews and physical examinations. All subjects provided written informed consent, and details of the DAYFUN protocol have been explained previously.^3,4^ The parent study protocol was approved by the NYU Institutional Review Board, and the secondary analysis of data from the parent study was approved by the Mount Sinai Institutional Review Board (IRB: STUDY-23-01038-AIMS). For the present study, a subset of healthy subjects with good PSG and SpO2 quality (n=16) was comprised in the analyses. All healthy subjects underwent full nocturnal polysomnography (NPSG) using the Sandman sleep system (Embla Systems Inc., Broomfield, CO, USA).

**Functional Imaging of Navigation across Sleep (FINS)**

The FINS study was a cross-sectional functional neuroimaging study designed to investigate the effects of sleep disruption induced by graded auditory stimuli and the underlying brain mechanisms correlated with spatial navigational memory performance.^5^ Healthy subjects without any no evidence of sleep disorders or OSA based on physician clinical interviews and in-lab PSG were recruited. All subjects provided written informed consent. For the present study, a subset of healthy subjects with good PSG and SpO_2_ quality (n=14) was included in the analyses. This study was approved by the Mount Sinai Institutional Review Board (IRB: FINS - STUDY-17-01313).

**Sleep Heart Health Study (SHHS)**

The SHHS was a multicenter prospective cohort study designed to investigate the association between OSA and cardiovascular and other clinical outcomes. Details of the study design have been described previously.^6^ Individuals aged ≥40 years without a history of treatment for sleep apnea, tracheostomy, or current home oxygen therapy were eligible for participation. A total of 6,441 subjects completed baseline assessments, including demographic questionnaires, sleep habit questionnaires, and overnight unattended in-home PSG. Institutional review board approval was obtained at all participating centers, and all subjects provided written informed consent. Of the 6,441 enrolled subjects, 637 withdrew consent because of sovereignty-related issues, resulting in a final SHHS dataset of 5,804 subjects. For the present study, only subjects with good-quality PSG and SpO₂ recordings (n=5,018) were included in the final analyses.

**Osteoporotic Fractures in Men (MrOS)**

The MrOS is multicenter prospective observational cohort study that enrolled 5,994 community-dwelling men aged ≥65 years from 6 sites across the United States. The study was originally designed to investigate the epidemiology of fractures in older men, and details of the study design have been described previously.^7-9^ All subjects provided written informed consent, and institutional review board approval was obtained at each participating center. Baseline assessments were conducted between 2000 and 2002, followed by a sleep examination between 2003 and 2005 that enrolled overnight unattended in-home PSG. For the present study, only subjects with good-quality PSG and SpO_2_ recordings were included in the final analyses (n = 2,759).

**Multi-Ethnic Study of Atherosclerosis (MESA)**

The MESA is a prospective population-based cohort study initiated in 2000 to investigate the prevalence, correlates, and progression of subclinical cardiovascular disease (CVD) in a diverse population of more than 6,000 men and women aged 45–84 years without known CVD at baseline. Subjects were recruited from 6 field centers across the United States. Institutional review board approval was obtained at all participating centers, and all subjects provided written informed consent. Details of the study design have been described previously.^10,11^ Overnight unattended in-home PSG was performed beginning in April 2010 during follow-up examination 11. A total of 1,544 subjects completed PSG assessments and sleep questionnaires with good-quality data. For the present study, only subjects with good-quality PSG and SpO₂ recordings were involved in the final analyses (n=1,326).

**Cleveland Family Study (CFS)**

The CFS is a longitudinal family-based cohort study established to investigate the genetic and familial determinants of sleep apnea.^12,13^ The cohort recruited families enriched for OSA from the Cleveland, Ohio area, including both African American and White subjects. Details of the study design have been described previously.^13^ Subjects underwent comprehensive clinical assessments and overnight PSG. Institutional review board approval was obtained, and all subjects provided written informed consent. A total of 566 subjects with good-quality PSG and SpO₂ recordings were comprised in the analyses.

**Wisconsin Sleep Cohort (WSC)**

The WSC is a longitudinal population-based cohort study established to investigate the natural history and health consequences of sleep disorders, particularly OSA.^14,15^ Subjects were recruited from state employees in Wisconsin and underwent repeated overnight in-lab PSG assessments along with detailed clinical and questionnaire evaluations. Details of the study design have been described previously.^14,15^ Institutional review board approval was obtained, and all subjects provided written informed consent. For the present study, 1,037 subjects were analyzed.

**The Apnea Positive Pressure Long-term Efficacy Study (APPLES)**

The APPLES was a multicenter, randomized, double-blind, sham-controlled clinical trial designed to evaluate the long-term effects of continuous positive airway pressure (CPAP) therapy on neurocognitive function, mood, sleepiness, and quality of life in patients with OSA.^16,17^ Subjects underwent comprehensive clinical assessments and overnight PSG across 5 centers in the United States. Details of the study design and procedures have been described previously.^16,17^ For the present study, 1,062 subjects were retained for analysis.

**Mount Sinai AI Research in Sleep Apnoea (AIRS) Cohort**

The AIRS cohort consists of individuals evaluated at sleep clinics across the Mount Sinai Health System between 2016 and 2023. Subjects were referred for in-laboratory PSG because of symptoms suggestive of sleep apnea and/or excessive daytime sleepiness. Inclusion criteria for the AIRS cohort were: (1) age >18 years, (2) suspected OSA, (3) suspected sleepiness, and (4) good-quality PSG signals with >5 hours of recording time. The final sample consisted of 2,263 subjects. This study was approved by the Mount Sinai Institutional Review Board (IRB: STUDY-23-01038-AIMS).
